# Association of the 609 C/T NAD(P)H: quinone oxidoreductase (NQO1) polymorphism with development of cutaneous malignant melanoma

**DOI:** 10.1101/2022.12.19.22283655

**Authors:** Maryam Naghynajadfard, Nikolas J Hodges, Adam Towler, Amanda J Lee, Joanna Norman, Ryan Doran, Rhiannon M David, Anne E Pheasant, Jerry Marsden, James K Chipman

## Abstract

Cutaneous Malignant Melanoma (CMM) is a life threatening disease whose incidence and mortality rates have risen rapidly in the White Caucasian population in recent decades. The aim of the current study was to investigate the association between polymorphisms in genes involved in DNA-repair and detoxification of reactive metabolites and the development of CMM. The patient cohort consisted of 69 individuals while the control population consisted of 100 individuals. We found a statistically significant association between the presence of the wild type NQO1 C allele, MDHFR C667T, TS*1494del6*, TSER polymorphisms and development of CMM [*P* = 0.04; odds ratio = 2.35]. The NQO1 CC genotype was more strongly associated with CMM development [*P* = 0.016; odds ratio = 2.92]. The NQO1 gene codes for a protein that has been widely considered to be protective through its ability to detoxify quinones. However recent studies have also linked it to an important source of reactive oxygen and to NF-κB-dependent proliferation of cultured melanoma cells. In conclusion these results link molecular epidemiology and experimental evidence for the role of the NQO1 gene product in development of CMM. MDHFR and TS in Folic acid metabolism are responsible for methylation of methyl group. Two important roles of folate ‘related to this study’ are the conversion of homocysteine to methionine and the generation of thymidylate (dTMP) which is required for DNA synthesis (Hayward, 2003). According to many studies done at this area, folate deficiency has been associated with chromosome strand breaks (Blount *et al*, 1997), impaired DNA repair (Hayward, 2003), DNA hypomethylation and hypermethylation all of which have been associated with cancer cell formation (Simonetta *et al* 2001) (Dong-Hyun K. 2007). The result of study shows, MTHFR C677T and TS 6bp deletion/insertion are not related to increased risk of CMM and therefore have no effect on an individual’s susceptibility.

## Introduction

Cutaneous Malignant Melanoma (CMM) is a life threatening disease whose incidence and mortality rates have risen rapidly in the White Caucasian population in recent decades. The consistant stimulation of cutaneous skin cells ‘melanocytes’ by weapons such as Biological mass destructions (WMD) which cause the change of skin colour to brown and hypertrophy, it calls cutaneous malignant melanoma (CMM) in some countries such as UK. However in some countries like Iran it calls Acne, it is removable by surgery and it is non lethal. It can be seen at the skin of face, neck, around the body and feet nail. There is considerable epidemiological evidence supporting a role of exposure to UV radiation as an important environmental factor in the aetiology of CMM. Interestingly intermittent sun exposure early in life rather than cumulative sun exposure appears to be a better indicator of disease risk (Zanetti *et al*, 1992, Fears *et al*, 2002). The strong aetiological role for UV exposure is clearly supported by the observation that childhood immigrants to Australia where ambient UV levels are amongst the highest in the world is associated with an increased lifetime risk of CMM. Individuals with light skin colour, freckles and a tendency to burn appear to be especially susceptible (Khlat *et al*, 1992, Green *et al*, 1988, reviewed by Jhappen *et al*, 2003). The observation that only a subset of individuals exposed to sunlight progress to develop CMM indicates that other factors including genetic variation may also play a role in the aetiology of CMM. In support of this, several high penetrance genes have being identified that are associated with risk of familial CMM these include mutations in genes encoding proteins involved in regulation of the cell cycle including CDK4 and the CDKIs p16 and p14ARF (reviewed by Hayword, 2003). However these mutations are rare and are only able to account for a relatively small percentage of cases of CMM.

UVB (290-320nm) is strongly absorbed by DNA and is known to induce a number of mutagenic DNA lesions including cyclobutane pyrimidine dimers and pyrimidine (6-4) photoproducts that are substrates for the nucleotide excision repair (NER) pathway in cells. In contrast the ability of UVA (320-400nm) to damage DNA has being largely ascribed to the generation of intracellular reactive oxygen species (ROS). For example UVA has being shown induce single stranded DNA breaks as well as 8-oxo 7,8-dihydro-2’-deoxyguanosine (8-oxo dG) (Besaratinia *et al* 2004, Cadet *et al*, 1997, Cooke *et al*, 2000, Douki *et al*, 1999, Kielbassa *et al*, 1997, Kvam *et al*, 1997, Zhang *et al*, 1997). These DNA lesions are repaired predominantly by the base excision repair pathway (BER).

Polymorphic genes involved in the NER and BER pathways and in the detoxification of ROS are therefore attractive candidates for genetic markers of individual susceptibility to CMM. Two key candidate genes in the BER pathway are OGG1, the major DNA glycosylase involved in the removal of 8-oxo dG from genomic DNA, and XRCC1. Interestingly, several studies have provided evidence that the cys326 variant of OGG1 has a reduced ability to restore repair activity in *E*.*Coli* deficient in the repair of 8-oxo dG (Kohno *et al*, 1998). In addition, the catalytic efficiency (k_cat_/k_M_) of excision of 8-oxo dG from γ-irradiated DNA by ser326 OGG1 protein is twice that of cys326 OGG1 (Audebert *et al*, 2000, Dherin *et al*, 1999). Moreover cys/cys genotype has being found to be a repair deficient phenotype (Chen *et al*, 2003, Aka *et al*, 2004, Lee *et al*, 2004). Although the precise role of XRCC1 remains unclear it is thought to play a structural role in the organisation of other BER repair factors (Thompson and West 2000). Recent studies have shown that the arg399gln polymorphism of XRCC1 is associated with reduced DNA repair capacity as assessed by the persistence of DNA adducts (Lunn *et al*, 1999) and strand breaks (Aka *et al*, 2004).

In addition, many of the enzymes believed to be involved in the detoxification of UVA-induced ROS (including GSTT1, GSTM1 and NQO1) are also polymorphic in the human population and as such are also attractive candidates for influencing individual cancer susceptibility. GSTM1 and GSTT1 null genotypes have been consistently associated with individual susceptibility to basal cell carcinoma (BCC) (reviewed by Lear *et al*, 2000) but their role in the aetiology of CMM remains far less clear. NQO1 is an obligate two-electron reductase involved in detoxification reactions converting quinones to hydroquinines (Ross *et al*, 2000) preventing single-electron redox cycling reactions. In addition NQO1 is believed to play a direct role in antioxidant defense by catalysing the reduction of α-tocopherolquinone to the antioxidant α-tocopherolhydroquinone (Ross *et al*, 2000). Interestingly the NQO1*0 allele which codes for an essentially non-functional protein (Traver *et al*, 1997) has also been identified as a candidate loci that is associated with individual susceptibility to BCC (Clairmont *et al*, 1999). However any possible role for this NQO1 polymorphism in susceptibility to CMM remains unexplored.

The aim of the current study was to investigate the possible association of a range of polymorphic genes involved in BER and the detoxification of ROS and individual susceptibility to CMM.

## Materials and Methods

### Study population

The patient cohort comprised 69 individuals with histologically confirmed malignant melanoma who were referred to the Skin Oncology Clinic, University Hospital, Birmingham NHS Trust. Clinical data for these individuals are presented in table 1. The control population consisted of 100 healthy volunteers with no family history of malignant melanoma. Because of the possible confounding effect of ethnicity all patients and volunteers were White Caucasians.

**Table 1:**
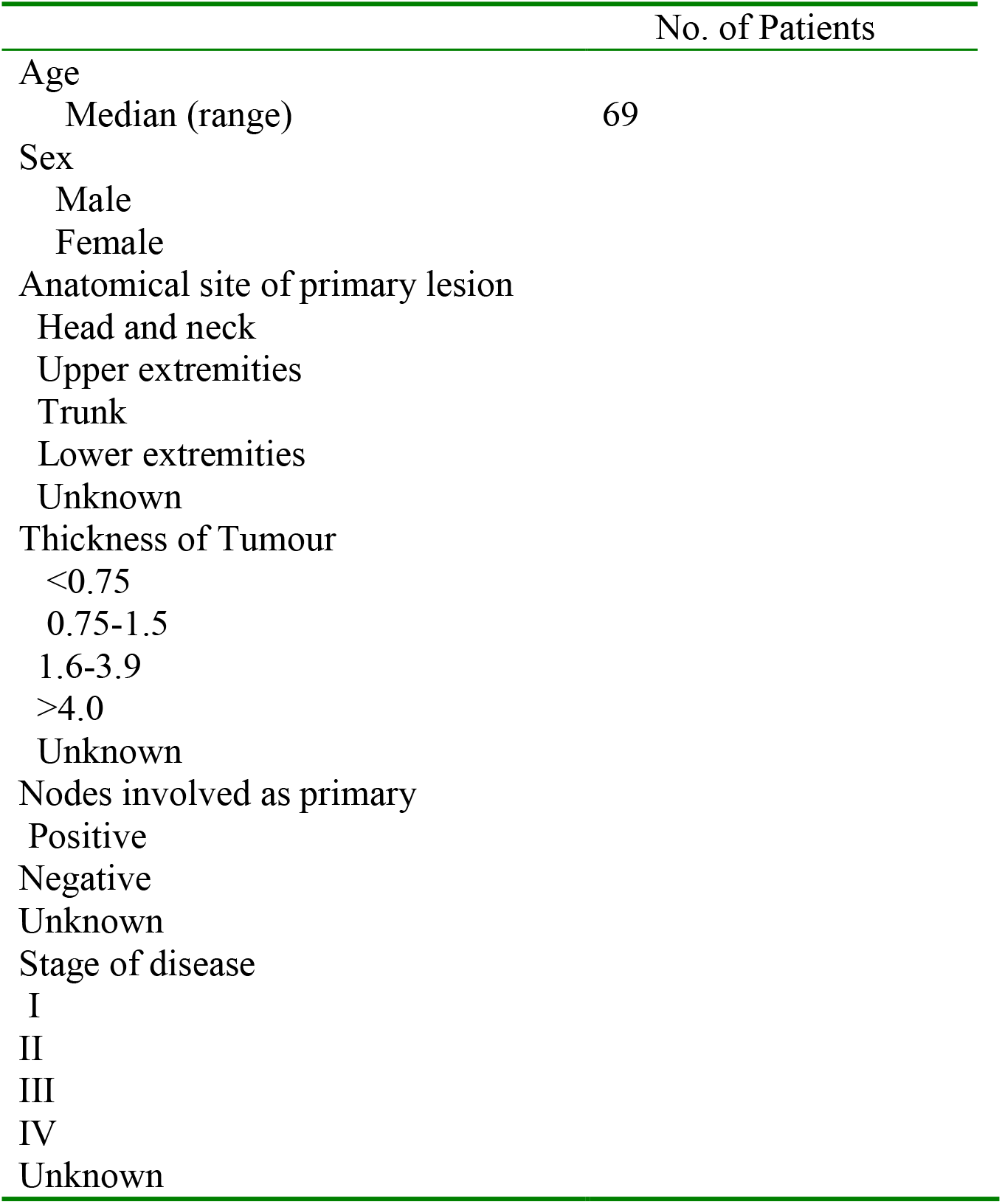
Clinical Data for malignant melanoma population.

|  | No. of Patients |
| --- | --- |
| Age |  |
| Median (range) | 69 |
| Sex |  |
| Male |  |
| Female |  |
| Anatomical site of primary lesion |  |
| Head and neck |  |
| Upper extremities |  |
| Trunk |  |
| Lower extremities |  |
| Unknown |  |
| Thickness of Tumour |  |
| <0.75 |  |
| 0.75-1.5 |  |
| 1.6-3.9 |  |
| >4.0 |  |
| Unknown |  |
| Nodes involved as primary |  |
| Positive |  |
| Negative |  |
| Unknown |  |
| Stage of disease |  |
| I |  |
| II |  |
| III |  |
| IV |  |
| Unknown |  |

### Blood Sample collection

Ethical approval was granted from South Birmingham (UK) Local Research Ethics Committee (reference number 0534) before commencement of the study. After written consent, blood samples (2ml) were obtained by venupuncture and collected in Vacutainer tubes (Haemograd, Becton-Dickinson, UK) containing EDTA. All samples were stored on ice until required and were processed within 3h of sample collection.

### Genotyping analyses

Genomic DNA was isolated from whole blood (approximately 1ml) using a QIAamp midi kit (Qiagen, UK), according to the manufacturer’s instructions. All primers were synthesised by AltaBiosciences (The University of Birmingham, UK). PCR genotyping reactions were carried out as previously reported to detect deletions in GSTM1 and GSTT1 (Arand *et al*, 1996), with the exception that GST and albumin (internal control) primers sequences were as detailed previously (Nair *et al*, 1999). In addition, previously reported RFLP-PCR methods were employed to detect common single nucleotide polymorphisms in NQO1, OGG1, XRCC1, XRCC3, ERCC2 (XPD), CYP2D6 and NAT2 (see Table 2 for details).

**Table 2:** Genes selected for PCR-RFLP analysis with position of single nucleotide polymorphisms and resulting amino acid change indicated.

| Gene<br>(Methodology reference) | Position of<br>polymorphism | Nucleotide<br>substitution | Amino acid<br>change |
| --- | --- | --- | --- |
| *NAT2*<br>Hickman and Sim (1991) | 341 | T→C | Ile<sup>114</sup> Thr (*NAT2* *5) |
|  | 590 | G→A | Arg<sup>197</sup> Gln (*NAT2* *6) |
|  | 857 | G→A | Gly<sup>286</sup> Glu (*NAT2* *7) |
| *CYP2D6*<br>Smith *et al*, (1992) | 1846 | G→A | Splicing defect<br>(*CYP2D6* *4) |
| *OGG1*<br>Hardie *et al*, (2000) | 1245 | C→T | Ser<sup>326</sup>Cys |
| *NQO1*<br>Traver *et al*, (1997) | 609 | C→T | Pro<sup>187</sup> Ser (*NQO1* *0) |
| *XRCC1*<br>Lunn *et al*, (1999) | 26304 | C→T | Arg<sup>194</sup>Trp |
|  | 28152 | G→A | Arg<sup>399</sup>Gln |
| *XRCC3*<br>De Ruyck *et al*, (2005) | 18067 | C→T | Thr<sup>241</sup>Met |
|  | 17 893 | A→G | IVS5-14 |
| *ERCC2 (XPD)*<br>Yu *et al* (2004) | 35931 | C→A | Lys<sup>751</sup>Gln |

### Statistical analysis

Phenotype frequencies were calculated by counting the number of individuals in a population positive for an allele (Σ [homozygotes + heterozygotes]). Allele frequencies were obtained directly by counting the number of chromosomes bearing an allele (Σ [homozygotes + 0.5x heterozygotes]). Associations were assessed using contingency table analysis and Fisher’s exact test. The OR and relative risk were calculated and a Bonferroni correction was applied to correct for multiple comparisons.

## Results

Phenotype, allele and genotype frequencies were calculated for the eleven single nucleotide polymorphisms (SNPs) examined (XRCC1 Arg^194^Trp, XRCC1 Arg^399^Gln, XRCC3 Thr^241^Met, XRCC3 IVS5-14, ERCC2 Lys^751^Gln, NQO1 Pro^187^Ser, OGG1 Ser^326^Cys, CYP2D6 G → A and NAT2). In the case of NAT2 Ile^114^Thr, Arg^197^Gln and Gly^286^Glu all represent alleles encoding a slow acetylator protein. These alleles were not distinguished in the analysis and were classified as slow (S) acetylator alleles. The wild type allele encodes a fast acetylator protein and was classified as a fast (F) allele. For analysis of GSTM and GSTT because our PCR test was not able to distinguish homozygous positive individuals from heterozygotes individuals were classified as either GST positive (wild type and heterozygous combined) or GST null. Analysis of the data revealed a statistically significant association with the C/T polymorphism at position 609 of NQO1 and the occurence of CMM. Individuals with a CC genotype had a significantly increased risk of developing CMM [OR=2.92 (1.38-6.1); *P* = 0.016 (Table 3). Presence of the C allele was also associated with development of CMM [OR=2.35 (1.23-4.5); *P* = 0.04. The association was further reduced in significance once the heterozygous genotype frequency was included and was not significantly different following application of the Bonferroni correction factor (*P* = 0.152). No associations with melanoma development were found with any of the other genetic polymorphisms studied (Table 3 and Table 4).

**Table 3:**
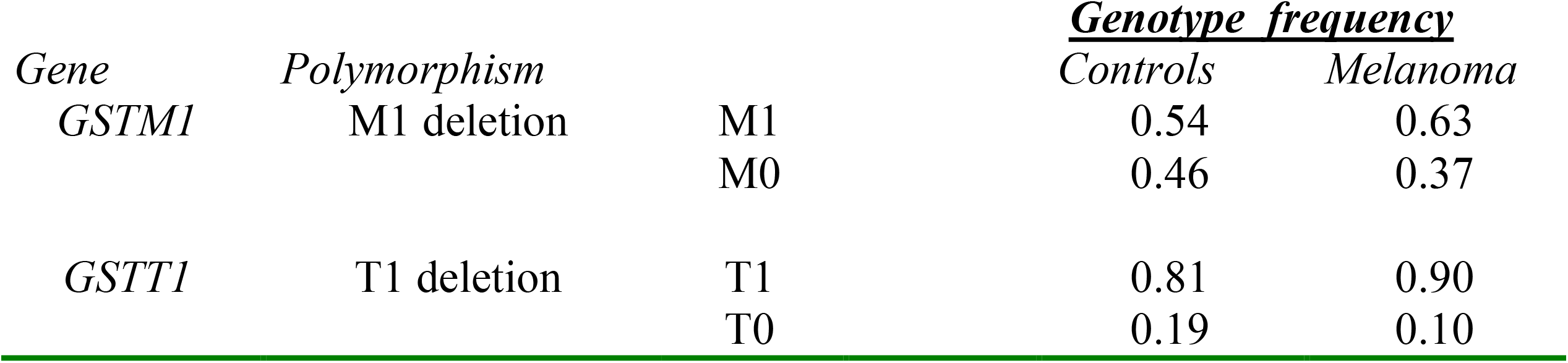
Analysis of GSTM1 and GSTT1 gene deletion frequencies in control and malignant melanoma cohorts.

**Table 4:**
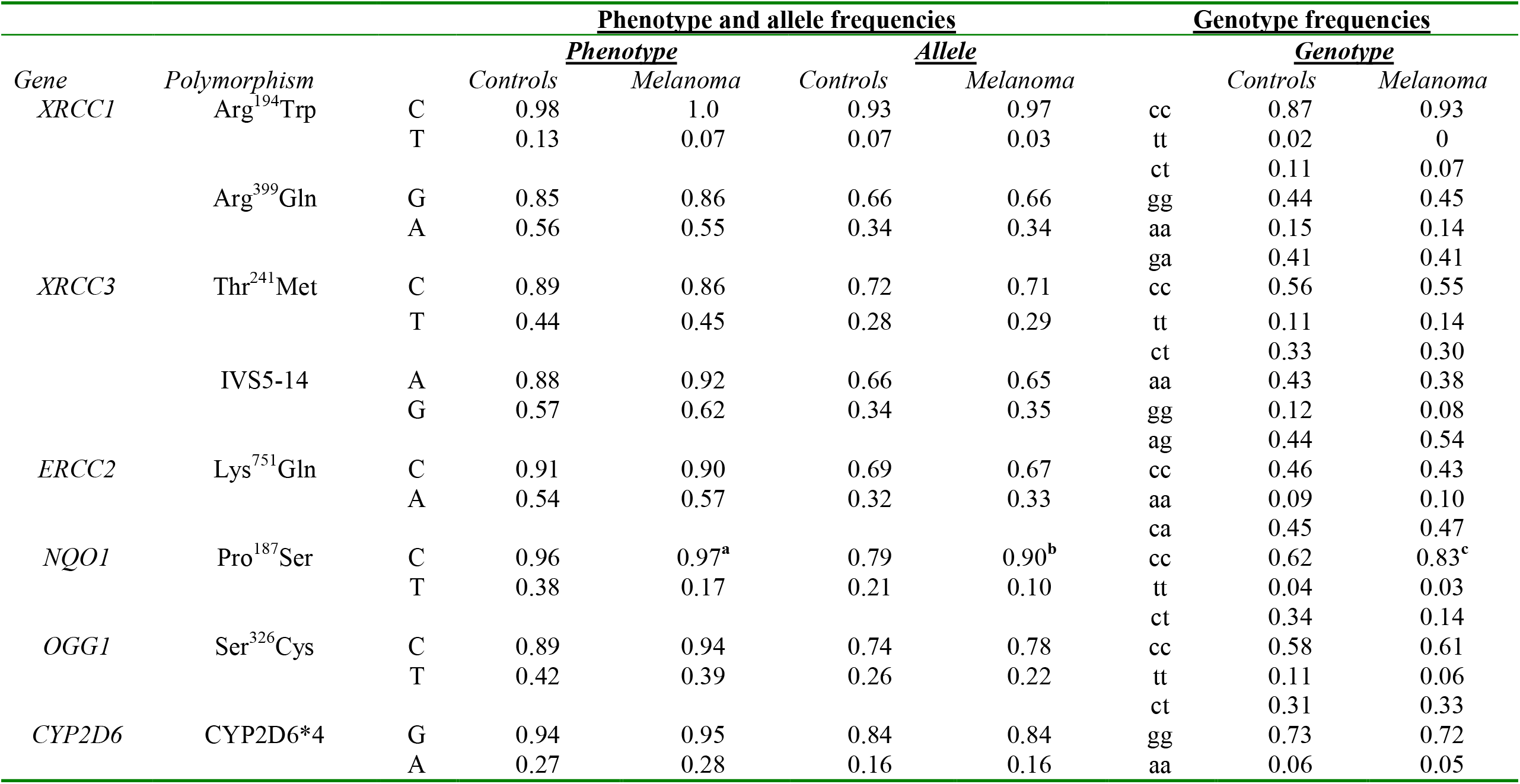
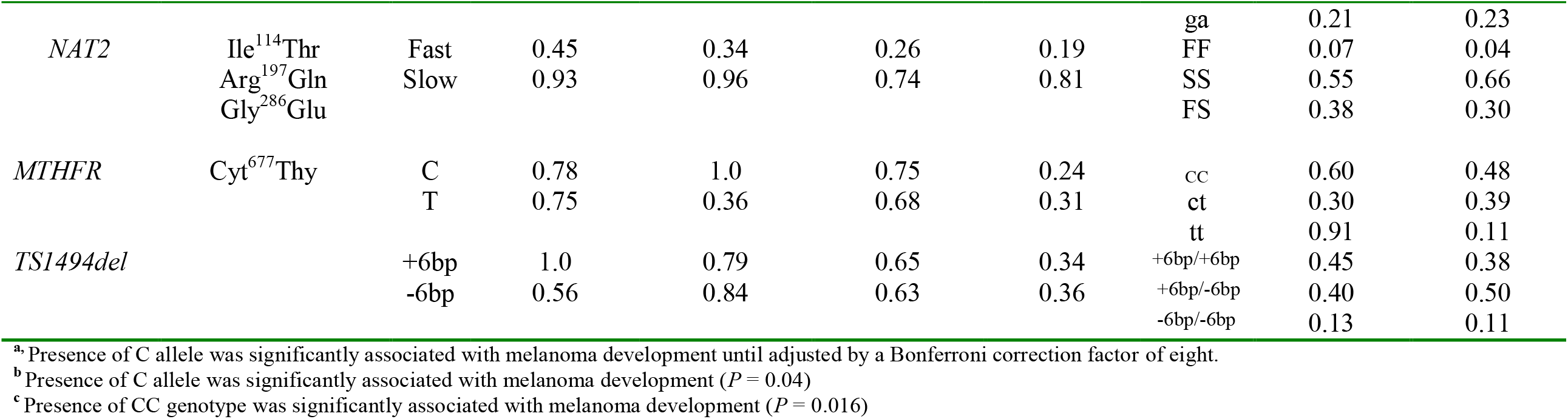
Phenotypic, allelic and genotype frequencies of gene polymorphisms in controls and malignant melanoma cohorts.

## Discussion

The aim of this study was to investigate whether genetic polymorphisms in a sample of genes whose products are involved in DNA repair and detoxification of reactive metabolites were associated with the development of CMM.

We found a statistically significant inverse association between an NQO1 polymorphic variant and risk of development of CMM (Table 3). NQO1 is believed to play a role in antioxidant defence and is able to catalyse the reduction of of α-tocopherolquinone to the antioxidant α-tocopherolhydroquinone (Ross *et al* 2000). The 609 C→ T (Pro^187^Ser) polymorphism was initially characterised in humans by Traver *et al* (1997). The Ser^187^ variant of NQO1 encodes a protein with essentially zero biological activity. This is believed to be related to a greatly enhanced rate of protein degradation of the variant form by the ubiquitin/proteosomal system (Ross *et al*, 2000). The frequency of the homozygous TT genotype has previously been reported to be approximately 5% in the Caucasian population (Kelsey *et al*, 1997). In agreement with this we observed a TT genotype frequency of 4.0% in our control population. Previous studies have associated the homozygous TT genotype with an elevated risk of a number of cancers (e.g. Benhamou *et al*, 2001, Menzel *et al*, 2004, Sarbia *et al*, 2003, Sunaga *et al*, 2002, Wiemels *et al*, 1999). Interestingly, Clairmont *et al*, (1999) reported that the NQO1*0 allele was a predictor of number of basal cell carcinomas in a multivariate model of patients of known GSTM1, GSTT1 and CYP2D6 EM genotype. However any possible role for this NQO1 polymorphism in susceptibility to CMM remains unexplored.

In the present study we observed that individuals with a “wild type” CC NQO1 genotype had a statistically significant increased risk of developing CMM [OR=2.92 (1.38-6.1); *P* = 0.016. Furthermore, presence of the C allele was also associated with development of CMM [OR=2.35 (1.23-4.5); *P* = 0.04.

Our findings are at first surprising in light of the apparent protective role of functionally active NQO1 protein that has previously been reported. However, NQO1 has also being reported to be greatly up regulated in tumours of the liver, lung, colon and breast (Belinsky *et al*, 1993). In particular, elevated levels of NQO1 enzyme activity in darkly pigmented congenital naevus cells and cultured melanoma cell lines has also being observed previously (Smit *et al*, 1999). It is possible that NQO1 activity may confer a possible growth/survival advantage to at least a subset of certain tumours.

Interestingly, Brar *et al*, (2001) report that constitutive activation of NF-κB is important for proliferation of in the malignant melanoma cell lines CRL1585 and CRL1619. NF-κB activation has also being reported to be important in supporting the survival and proliferation of a number of cancers including carcinomas of the breast, ovary, colon, lung, head and neck and pancreas (Batra *et al*, 1999, Bours *et al*, 1994, Duffey *et al*, 1999, Sovak *et al*, 1997). NF-κB is constitutively activated in melanoma cell lines (Meyskens *et al*, 1999) including the Hs294T line, where activation was observed as a result of enhanced degradation of the inhibitory partner inhibitory κB alpha (IκBα) (Shattuck-Brandt & Richmond, 1997). No such activation is observed in normal melanocytes (McNulty *et al*, 2004, Meyskens *et al*, 1999) In the study by Brar *et al* (2001) NF-κB activation and cell growth was inhibited by antioxidants suggesting an intermediary role of ROS. In addition, an inhibitor of NQO1 (dicumarol) inhibited NF-κB activation and cellular growth indicating a possible role of NQO1 in ROS generation in these cell lines. Based on results using a quinone analogue (capsaicin) the authors speculated that the source of ROS may involve a NQO1/quinone redox couple. Elevated levels of ROS has also been observed in dysplastic naevi compared to normal skin melanocytes (Pavel *et al*, 2004). Dysplastic naevi are widely considered to be melanoma precursors further supporting the hypothesis that aberrant redox homeostasis is important in melanoma development.

It must be noted however, that other sources of ROS are also considered to contribute to NF-κB activation in melanoma cell lines. These include NAD(P)H oxidase activity (Brar *et al* 2002) and gamma-glutamyl transferase (GGT) (Dominici *et al* 2003, Pieri *et al*, 2003). Indeed the inhibitory effects of dicurmarol on NFκB activation in melanoma cells appears to be at least in part, to be due to inhibition of NAD(P)H oxidase (Brar et al 2002). Morre *et al*, (1995, 1996) have also shown a similar inhibition of growth of human and mouse tumour lines including those derived from melanoma by capsaicin but this was considered to be related to inhibition of NAD(P)H oxidase activity. NQO1 may nevertheless contribute to the survival and proliferation advantage as discussed above. A pro-oxidant role of melanin has being proposed in the pathogenesis of melanoma (reviewed by Meyskens *et al* 2001) and it is possible that NQO1 could also generate ROS by promoting redox cycling of melanin.

Together these studies indicate that intracellular ROS from multiple sources contribute to melanoma cell survival and growth. We hypothesise that under certain circumstances the wild type NQO1 gene product contributes to the promotion of growth of cutaneous malignant melanomas *in vivo* either by the mechanism proposed by Brar *et al* (2001) or by promoting redox cycling of melanin. This biologically plausible mechanism may also explain our observation that the wild type NQO1 C allele and NQO1 CC genotype is significantly associated with melanoma development.

This study used a candidate gene approach to analyse genetic factors involved in disease susceptibility. A major potential problem in this approach is the possibility of finding associations by chance. We have addressed this by applying the conservative Bonferroni correction for multiple comparisons and by only considering an association significant if it maintained a *P* < 0.05 following correction. Although the data presented in this study indicate a role for NQO1 in individual susceptibility to CMM further work is required with larger sample numbers to confirm this association. In addition, work to investigate levels of ROS in NQO1 wild type and mutant melanoma cells is planned.

In conclusion, this study has identified a genetic factor that may influence an individual’s susceptibility to CMM. The data presented also provide further insight into the possible biological role of NQO1 in CMM tumour progression. Antioxidants have been shown to inhibit both growth in both melanoma cell lines and tumours in animal models (e.g. Calagirone *et al*, 2000, Tanaka *et al*, 2000, Liu *et al*, 2001, Malafa *et al*, 2002 Kogure *et al*, 2003, reviewed by Sander *et al*, 2004). Therefore, NQO1 wild type individuals and those with elevated NAD(P)H oxidase may represent prime candidates for the future testing of novel therapeutic strategies involving the use of antioxidants for interruption of oxidant signalling pathways involved in melanoma growth.

## Data Availability

All data produced in the present study are available upon reasonable request to the authors
All data produced in the present work are contained in the manuscript

## Abbreviations used

(CMM): Cutaneous Malignant Melanoma
(UV): Ultraviolet
(CDK): cyclin dependent kinase
(CDKI): cyclin dependent kinase inhibitor
(ROS): reactive oxygen species
(8-oxo dG): 8-oxo 7,8-dihydro-2’-deoxyguanosine
(OGG1): 8 oxo deoxyguanosine DNA glycosylase 1
(NAT): N-acetyl transferase
(NQO1): NAD(P)H: quinone oxidoreductase
(GST): glutathione-S-transferase
(XRCC1): X-ray repair complementing defective repair in Chinese hamster cells 1

## Acknowledgements

The authors wish to acknowledge ICI, The European Social Fund and the COLT foundation for funding that has made this study possible.

## Notes

### Competing Interest Statement

The authors have declared no competing interest.

### Funding Statement

Birmingham university and personal funding

### Author Declarations

Ethical approval was granted from South Birmingham (UK) Local Research Ethics Committee (reference number 0534) before commencement of the study.

## Bibliography

Aka P, Mateuca R, Buchet J-P, Thierens H and Kirsch-Volders M. Are genetic polymorphisms in OGG1, XRCC1 and XRCC3 predictive for the DNA strand repair phenotype and genotoxicity in workers exposed to low dose ionising radiations? Mutat Res. 2004; 556: 169–81.

Arand M, Muhlbauer R, Hengstler J, Jager E, Fuchs J, Winkler L. Multiplex polymerase chain reaction protocol for the simultaneous analysis of the glutathione S-transferase GSTM1 and GSTT1 polymorphisms. Annal Biochem. 1996; 236: 184–86.

Audebert M, Radicella JP. and Dizdaroglu M. Effect of single mutations in the OGG1 gene found in in human tumours on the substrate specificity of the OGG1 protein. Nuc. Acids. Res.2000; 28: 2672–78.

Batra RK, Guttridge DC, Brenner DA, Dubinett SM, Baldwin AS, Boucher RC. IkappaBalpha gene transfer is cytotoxic to squamous-cell lung cancer cells and sensitizes them to tumor necrosis factor-alpha-mediated cell death. Am J Respir Cell Mol Biol. 1999; 21:238–45.

Belinsky M, Jaiswal AK. NAD(P)H:quinone oxidoreductase1 (DT-diaphorase) expression in normal and tumor tissues. Cancer Metastasis Rev. 1993;12:103–17.

Benhamou S, Voho A, Bouchardy C, Mitrunen K, Dayer P. and Hirvonen A. Role of NAD(P)H quinoneoxidoreductase polymorphism at codon 187 in susceptibility to lung, laryngeal and oral/pharyngeal cancers. Biomarkers 2001; 66: 440–7.

Besaratinia A, Synold TW, Bixin X and Pfeifer GP. G-to-T transversions and small tandem base deletions are the hallmark of mutations induced by ultraviolet A radiation in mammalian cells. Biochemistry 2004; 43: 8169–77.

Bours V, Dejardin E, Goujon-Letawe F, Merville MP, Castronovo V. The NF-kappa B transcription factor and cancer: high expression of NF-kappa B- and I kappa B-related proteins in tumor cell lines. Biochem Pharmacol. 1994; 47: 145–9.

Brar S, Kennedy TP, Whorton R, Sturrock AB, Huecksteadt TP, Ghio AJ. et al. Reactive oxygen species from NAD(P)H quinone oxidoreductase constitutively activate NF-kB in malignant melanoma cells. Am. J. Physiol Cell Physiol. 2001; C659–76.

Brar SS, Kennedy TP, Sturrock AB, Huecksteadt TP, Quinn MT, Whorton AR, Hoidal JR. An NAD(P)H oxidase regulates growth and transcription in melanoma cells. Am J Physiol Cell Physiol. 2002 Jun;282:C1212–24.

Cadet J, Berger M, Douki T, Morin B, Raoul S, Ravanat J. and Spinelli S. Effects of UV and visible radiation on DNA-final base damage. Biol. Chem. 1997; 378: 1275–86.

Caltagirone S, Rossi C, Poggi A, Ranelletti FO, Natali PG, Brunetti M, Aiello FB. and Piantelli M. Flavonoids apigenin and quercetin inhibit melanoma growth and metastatic potential. Int J Cancer. 2000; 87: p595–600.

Chen S-K., Hsieh WA, Tsai M-H, Chen C-C, Hong AI, Wei Y-Y. and Chang WP. Age-associated decrease of oxidative repair enzymes, human 8-oxoguanine DNA glycosylase I (hOGG1) in human ageing. J. Rad. Res. 2003; 44: 31–5.

Clairmont A, Sies H, Ramachandran S, Lear JT, Smith AG, Bowers B. et al. Association of NAD(P)H:quinone oxidoreductase (NQO1) null with the numbers of basal cell carcinomas: use of a multivariate model to rank the relative importance of this polymorphism and those at other relevant loci. Carcinogenesis 1999; 20: 1235–40.

Cooke MS, Mistry N, Ladapo A, Herbert KE. and Lunec J. Immunochemical quantification of UV-induced oxidative and dimeric DNA damage to human keratinocytes. Free Rad Res. 2000; 33: 369–81.

De Ruyck, K. et al., Radiation-induced damage to normal tissues after radiotherapy in patients treated for gynaecologic tumors: association with single nucleotide polymorphisms in XRCC1, XRCC3, and OGG1 genes and in vitro chromosomal radiosensitivity in lymphocytes, Int. J. Radiation Oncology Biol. Phys. Xx (2005) xx–xx

Douki T, Perdiz D, Grof P, Kuluncsics Z, Moustacchi E, Cadet J and Sage E. Oxidation of guanine in cellular DNA by solar radiation : biological role. Photochem Photobiol. 1999; 70: 184–90.

Dominici S, Visvikis A, Pieri L, Paolicchi A, Valentini MA, Comporti M, Pompella A. Redox modulation of NF-kappaB nuclear translocation and DNA binding in metastatic melanoma. The role of endogenous and gamma-glutamyl transferase-dependent oxidative stress. Tumori. 2003; 89: 426–33.

Dherin C, Radicella JP, Dizdaroglu M. and Boiteux, S. Excision of oxidatively damaged DNA bases by the human alpha –hOgg1 protein and and the polymorphic alpha-hOgg1 (ser326cys) protein which is frequently found in human populations. Nuc. Acids. Res.1999; 27: 4001–7.

Duffey DC, Chen Z, Dong G, Ondrey FG, Wolf JS, Brown K, Siebenlist U, Van Waes C. Expression of a dominant-negative mutant inhibitor-kappaBalpha of nuclear factor-kappaB in human head and neck squamous cell carcinoma inhibits survival, proinflammatory cytokine expression, and tumor growth in vivo. Cancer Res. 1999; 59: 3468–74.

Fears TR, Bird CC, Guerry D, Sagebiel RW, Gail MH, Elder DE et al. Average midrange ultraviolet radiation flux and time outdoors predict melanoma risk. Cancer Res. 2002; 62: 3992–6.

Green A, Sorahan T, Pope D, Siskind V, Hansen M, Hanson L. et al. Moles in Australian and British school children. Lancet 1988; 2: 1497.

Hardie LJ, Briggs JA, Davidson LA, Allan JM, King RFGJ, Williams GI, et al. The effect of OGG1 and glutathione peroxidase I genotypes and 3p chromosomal loss on 8-hydroxydeoxyguanosine levels in lung cancer. Carcinogenesis. 2000; 21: 167–72.

Hayword N. Genetics of melanoma predisposition. Oncogene. 2003; 22: 3053–62.

Hickman D, Sim E. N-acetyltransferase polymorphism: Comparison of phenotype and genotype in humans. Biochem. Pharmacol. 1991; 42: 1007–14.

Jhappan C, Noonan, Fp. and Merlino G. Ultraviolet radiation and cutaneous melanoma. Oncogene 2003; 22: 3099–3112.

Khlat M, Vail A, Parkin M, Green A. Mortality rates from melanoma in migrants to Australia: variation by age at arrival and duration of stay. Am J Epidemiol. 1992; 135: 1103–13.

Kelsey KT, Wiencke JK, Christiani DC, Zuo Z, Spitz MR, Xu X. et al. Ethnic variation and prevalence of a common NAP(H)quinone oxidoreductase polymorphism and its implications for anticancer chemotherapy. British Journal of Cancer. 1997; 76: 853–54.

Kielbassa C, Roza L. and Epe B. Wavelength dependence of oxidative DNA base damage induced by UV and visible light. Carcinogenesis. 1997; 18: 2379–84.

Kogure K, Manabe S, Hama S, Tokumura A. and Fukuzawa K. Potentiation of anti-cancer effect by intravenous administration of vesiculated alpha-tocopheryl hemisuccinate on mouse melanoma in vivo. Cancer Lett. 2003; 192: 19–24

Kohno T, Shinmura K, Tasaka M Tani M, Kim SR, Sugimura H et al. Genetic polymorphisms and alternative splicing of the hOGG1 gene that is involved in the repair of 8-hydroxyguanine in damaged cells. Oncogene 1998; 16: 3219–25.

Kvam E. and Tyrell RM. Induction of oxidative DNA base damage in human skin cells by UV and near visible radiation. Carcinogenesis. 1997; 18: 2379–84.

Lear JT, Smith AG, Strange RC and Fryer AA. Detoxifying enzyme genotypes and susceptibility to cutaneous malignancy. British Journal of Dermatology 2000; 142: 8–15.

Lee AJ, Hodges NJ and Chipman JK. Inter-individual variability in response to sodium dichromate induced oxidative DNA damage: Role of the Ser326Cys polymorphism in the DNA-repair protein of 8-oxo 7,8-dihydro-2’-deoxyguanosine DNA glycosylase 1 (OGG1). Cancer Epidemiology Biomarkers and Prevention. 2004; In Press

Liu JD, Chen SH, Lin CL, Tsai SH. and Liang YC. Inhibition of melanoma growth and metastasis by combination with (-)-epigallocatechin-3-gallate and dacarbazine in mice. J Cell Biochem 2001; 83: 631–42.

Lunn RM, Langlois RG, Gsieh LL, Thompson CL, Bell DA. XRCC1 polymorphisms: effects on aflatoxin B1-DNA adducts and glycophorin A variant frequency. Cancer Res. 1999; 59: 2557–61.

Malafa MP, Fokum FD, Mowlavi A, Abusief M. and King M. Vitamin E inhibits melanoma growth in mice. Surgery 2002; 131: 85–91.

McNulty SE, del Rosario R, Cen D, Meyskens FL Jr, Yang S. Comparative expression of NFkappaB proteins in melanocytes of normal skin vs. benign intradermal naevus and human metastatic melanoma biopsies. Pigment Cell Res. 2004; 17: 173–80.

Menzel HJ, Sarmanova J, Soucek P, Berberich R, Grunewald K, Haun M, Kraft HG. Association of NQO1 polymorphism with spontaneous breast cancer in two independent populations. Br J Cancer. 2004; 90:1989–94.

Morre DJ, Chueh PJ, Morre DM. Capsaicin inhibits preferentially the NADH oxidase and growth of transformed cells in culture. Proc Natl Acad Sci U S A. 1995; 92:1831–5.

Morre DJ, Sun E, Geilen C, Wu LY, de Cabo R, Krasagakis K, Orfanos CE, Morre DM. Capsaicin inhibits plasma membrane NADH oxidase and growth of human and mouse melanoma lines. Eur J Cancer. 1996; 32A:1995–2003.

Meyskens FL Jr, Farmer P, Fruehauf JP. Redox regulation in human melanocytes and melanoma. Pigment Cell Res. 2001; 14:148–54.

Meyskens FJ Jr, Buckmeier JA, McNulty SE and Tohidian NF. Activation of nuclear factor-kB in human metastic melanoma cells and effects of oxidative stress. Clin Can Res. 1999; 5: 1197–1202.

Nair UJ, Nair J, Mathew B, Bartsch H. Glutathione S-transferase M1 and T1 null genotypes as risk factors for oral leukoplakia in ethnic Indian betal quid/tobacco chewers. Carcinogenesis. 1999; 20: 743–48.

Pavel S, van Nieuwpoort F, van der Meulen H, Out C, Pizinger K, Cetkovska P, Smit NP, Koerten HK. Disturbed melanin synthesis and chronic oxidative stress in dysplastic naevi. Eur J Cancer. 2004 Jun; 40:1423–30.

Pieri L, Dominici S, Del Bello B, Maellaro E, Comporti M, Paolicchi A, Pompella A. Redox modulation of protein kinase/phosphatase balance in melanoma cells: the role of endogenous and gamma-glutamyltransferase-dependent H2O2 production. Biochim Biophys Acta. 2003;1621:76–83.

Ross D, Kepa JK, Winski SL, Beall HD, Anwar A. and Seagal D. NAD(P)H quinone oxidoreductase 1 (NQO1): chemoprotection, bioactivation, gene regulation and genetic polymorphisms. Chemico-Biological Interactions 2000; 129: 77–97.

Sanders CS, Chang H, Hamm F, Elsner P. and Thicle JJ. Role of oxidative stress and the antioxidant network in cutaneous carcinogenesis. Int J Dermatol 2004; 43: 326–35.

Sarbia M, Bitzer M, Siegel D, Ross D, Schulz WA, Zotz RB, Kiel S, Geddert H, Kandemir Y, Walter A, Willers R, Gabbert HE. Association between NAD(P)H: quinone oxidoreductase 1 (NQO1) inactivating C609T polymorphism and adenocarcinoma of the upper gastrointestinal tract. Int J Cancer. 2003; 10: 381–6.

Shattuck-Brandt RL and Richmond A. Enhanced degredation of IκBα contributes to endogenous activation of NF-κB in Hs294T melanoma cells. Cancer Research 1997; 57: 3032–39.

Smith CAD, Gough AC, Leigh PN, Summers BA, Harding AE, Maranganore, DM, et al. Debrisoquine hydroxylase gene polymorphism and susceptibility to Parkinson’s disease. The Lancet. 1992; 339: 1375–77.

Smit NP, Hoogduijn MJ, Riley PA, Pavel S. Study of DT-diaphorase in pigment-producing cells. Cell Mol Biol. 1999; 45: 1041–6.

Sovak MA, Bellas RE, Kim DW, Zanieski GJ, Rogers AE, Traish AM, Sonenshein GE. Aberrant nuclear factor-kappaB/Rel expression and the pathogenesis of breast cancer. J Clin Invest. 1997; 100: 2952–60.

Sunanga N, Kohno T, Yanagitani N, Sugimura H, Kunitoh H, Tamura T. et al. Contribution of the NQO1 and GSTT1 polymorphisms to lung adenocarcinoma susceptibility. Cancer Epidemiology, Biomarkers and Prevention. 2002; 11: 730–8.

Tanaka T, Kohno H, Murakami M, Kagami S. and El-Bayoumy K. Suppressing effects of dietary supplementation of the organoselenium 1,4-phenylenebis(methylene)selenocyanate and the citrus antioxidant auraptene on lung metastasis of melanoma cells in mice. Cancer Res. 2000; 60: 3713–6.

Thompson LH and West MG. XRCC1 keeps DNA from getting stranded. Mutat Res. 2000; 459: 1–18.

Traver RD, Siegel D, Beall HD, Phillips RM, Gibson NW, Franklin WA, et al. Characterization of a polymorphism in NAD(P)H: quinone oxidoreductase (DT-diaphorase). Br. J. Cancer. 1997; 75: 69–75.

Wiemels JL, Pagnamenta A, Taylor GM, Eden OB, Alexander FE and Greaves MF. A lack of a functional NAD(P)H quinone oxidoreductase allele is selectively associated with pediatric leukemias that have MLL fusions. Cancer Res 1999; 59: 4095–99.

Zanetti R, Franceschi S, Rosso S, Colonna S and Bidoli. Cutaneous melanoma and sunburns in childhood in a Southern European population. Eur J Cancer. 1992; 28A: 1172–6.

Zhang X, Rosenstein BS, Wang Y, Lebwohl M, Mitchel DM. and Wei H. Induction of 8-oxo 7,8-dihydro-2’-deoxyguanosine by ultraviolet radiation in calf thymus DNA 41and HeLa cells. Photochem Photobiol. 1997; 65: 119–24.

Yu, H.-P. et al., Polymorphisms in the DNA repair gene XPD and susceptibility to esophageal squamous cell carcinoma, Cancer Genet. Cytogenet. 154 (2004) 10-15.0

